# Cardiovascular outcomes in adult patients with atrial septal defect: a nationwide population-based study

**DOI:** 10.1101/2024.06.06.24308567

**Authors:** Jue Seong Lee, Man Young Park, Jin-Man Jung, Hong Ju Shin

## Abstract

**Background:** The beneficial effects of atrial septal defect (ASD) closure on cardiovascular events and mortality in adults remain undetermined. Therefore, this study aimed to investigate the long-term effects of different ASD closure methods on cardiovascular events in adults.

**Methods:** A retrospective analysis was conducted using data obtained from the Korean National Health Insurance Service from 2002 to 2020, focusing on patients aged ≥20 years diagnosed with ASD based on the codes provided by the Tenth revision of International Classification of Diseases codes between 2004 and 2015. Participants were categorized into the observation, device closure, and surgery groups. Propensity score matching (PSM) of variables such as sex, age, comorbidities, and medications was employed at a 2:1:1 ratio to mitigate imbalances among the groups. The Cox proportional hazards model was utilized to compare the occurrence of major adverse cardiovascular events (MACE), including stroke, myocardial infarction (MI), coronary revascularization, and all-cause death, and each individual component among the three groups.

**Results:** In total, 20,643 patients with ASD were included in this study. After PSM, there were 6,636 in the observation group and 3,318 each in the device closure and surgery group. Over a 5-year follow-up period, the adjusted hazard ratios for MACE were significantly lower in the surgery (0.72; 95% CI: 0.66–0.79) and device closure groups (0.85; 95% CI: 0.78–0.92) than in the observation group. Beneficial effects on stroke and all-cause mortality were observed in both intervention groups. Additionally, a beneficial effect on coronary revascularization was observed in the surgery group, whereas the impact on MI was not significantly different between the groups.

**Conclusions:** ASD closure, whether by surgery or using a device, is associated with a decreased incidence of cardiovascular outcomes in adults. The benefits on cardiovascular outcomes vary with the type of closure method, underscoring the need for a tailored approach to manage ASD in adults.

**Clinical Perspectives:** **What is new?**

- Compared with no intervention, surgery exhibited a more beneficial effect in terms of MACE, followed by device closure.
- Device closure appears to be more beneficial in terms of all-cause death.
- Surgery reduced the occurrence of coronary revascularization.

**What are the clinical implications?**

- Cardiovascular preventive effects after ASD closure (through surgery or device) were observed in adult patients with ASD when compared to the untreated individuals.
- The benefits on cardiovascular outcomes vary with the type of closure method, underscoring the need for a tailored approach to manage ASD in adults.

## Introduction

Atrial septal defect (ASD) is one of the most common congenital heart defects, and individuals with ASD often remain asymptomatic until adulthood.^1,2^ Surgical procedures and device closure have served as corrective treatments for ASD. Furthermore, with the widespread adoption of early diagnostic and treatment approaches, the mortality rate in patients who undergo early ASD closure is low, and long-term outcomes are estimated to be highly favorable.^1,3–5^ However, if not promptly treated, the risk of complications such as heart failure, stroke, and mortality increases.^1,6,7^ Even when ASD closure is performed in a timely manner, individuals with ASD may have higher rates of cardiovascular morbidity, mortality, and conditions such as ischemic heart disease, stroke, and atrial fibrillation (AF), compared with the general population.^2,8–10^

Currently, device closure is generally preferred over surgery for secundum ASD, while surgical intervention remains the preferred approach for other types of ASD.^1,4^ In a study comparing the outcomes of device closure and surgery in ASD treatment, mortality rates were not significantly different between the two treatment methods.^9^ Although some studies have reported a lower risk of AF in cases where device closure is performed, one particular study did not report any significant difference.^8,11^ The impact of ASD on cardiovascular outcomes may vary with age.^1^ Additionally, the age at which ASD closure is performed may also influence the outcome.^3^ In particular, adult patients with ASD who did not undergo closure during childhood are estimated to have a significantly higher risk of cardiovascular complications as they age. However, there remains a lack of research on how outcomes vary following ASD closure and whether different treatment methods, such as device closure or surgery, differentially influence these outcomes.

This study aimed to compare composite cardiovascular outcomes, including stroke, myocardial infarction (MI), coronary revascularization, and mortality, as well as each individual component of these composite outcomes between untreated adult patients with ASD and those who underwent surgery or device-based closure, using nationwide cohort data. Additionally, this study aimed to compare post-procedural cardiovascular outcomes until 6 months after ASD closure, specifically in patients who underwent surgery or device closure.

## Methods

### Study population

We retrospectively reviewed the data from the Korean National Health Insurance Service (NHIS) claims database. The Korean NHIS covers 98% of the South Korean population and provides a dataset that incorporates diagnostic codes according to the Tenth Revision of the International Classification of Diseases (ICD-10).^12^ It also includes demographic information, such as sex, age, economic status, and residential location, as well as comprehensive medical details encompassing prescriptions, examinations, interventions, and procedures carried out during hospitalizations and outpatient visits.

We initially extracted the data of adult patients aged ≥20 years diagnosed with ASD (Q211X) based on ICD-10 codes between 2002 and 2020 (**Supplementary Table 1**). Subsequently, we excluded patients with persistent foramen ovale (Q2110). To exclude patients diagnosed before 2002, a washout period of 2 years was implemented, thereby excluding individuals diagnosed between 2002 and 2003. Additionally, patients diagnosed between 2016 and 2020 were excluded to ensure a minimum follow-up period of 5 years. We also excluded all patients diagnosed with other congenital heart diseases regardless of the presence of ASD and those who experienced cardiovascular events before the ASD index date. For the observation group, the reference point was the time of diagnosis, whereas for the surgery and device closure groups, the index date was defined as the time of the procedure.

We segregated the study population into three groups: observation, surgery, and device closure.

### Variables and comorbidities

We investigated underlying conditions, such as hypertension (HTN), diabetes, pulmonary HTN, congestive heart failure (CHF), and endocarditis, in patients diagnosed with ASD (based on ICD-10 codes) (**Supplementary Table 1**). HTN and diabetes were defined according to both an ICD-10 code and a prescription for the respective medication. The use of statins, antiplatelet agents, and anticoagulants was also examined.

### Outcomes of interest

We investigated the occurrence of cardiovascular events following the ASD index date in each group during the 5-year follow-up period. The primary outcome of interest was major adverse cardiovascular events (MACEs) as a composite outcome including MI or any stroke coincident with hospitalization, coronary revascularization, and all-cause death. The secondary outcomes were the individual components of the composite outcome. The ICD-10 codes are provided in **Supplementary Table 1**. Furthermore, to assess post-procedural outcomes, we investigated the differences in the occurrence of cardiovascular outcomes within 6 months of the procedures between the surgery and device closure groups.

This study was approved by the Institutional Review Board of Korea University Ansan Hospital (AS0162). Informed consent was waived due to the retrospective nature of the study. To ensure privacy protection, participant identification numbers in the database were de-identified and encrypted. Therefore, the requirement of informed consent was waived.

### Statistical analysis

We implemented a propensity score matching (PSM) technique to address the observed disparities between the control and intervention groups, particularly regarding variables such as age, sex, HTN, diabetes, CHF, pulmonary HTN, and medications. This approach was essential for enhancing group comparability by equalizing the distribution of biases and confounding variables, thereby facilitating a more accurate estimation of intervention effects in observational studies similar to this one. Propensity scores representing the conditional probability of receiving treatment based on observed covariates were calculated using logistic regression models. In this study, three patient groups needed to be matched; therefore, we performed a 2:1:1 matching to maximize the number of participants.

Continuous variables were presented as mean ± standard deviation. Categorical variables were presented as counts and percentages. Baseline characteristics were compared using the Wilcoxon signed-rank test for continuous variables and the chi-square test for categorical variables. PSM was performed with a ratio of 2:1:1 for the observation, surgery, and device closure groups. Cox proportional hazard models were used to compare event rates between the three groups in terms of primary and secondary outcomes. Variables with limited numbers that could not be matched were further adjusted as covariates. Regarding MACE, a subgroup analysis was conducted to investigate the differences in outcomes based on the presence or absence of each risk factor. Statistical analyses were conducted using SAS software (version 7.1; SAS Institute, Cary, NC, USA).

## Results

A total of 20,643 patients with ASD were included in the study, following which 13,103 were included in the observation group. Surgery was performed in the case of 4,222 patients, and device closure was performed in 3,318 patients. The mean age was 45.4 ± 15.5 years, with males accounting for 6,967 individuals (33.7%).

After PSM, the final study population consisted of 6,636 patients in the observation group and 3,318 patients each in the surgery and device closure group. A flowchart of the study is shown in **Figure 1**. No statistically significant differences were observed for the following variables: sex, HTN, endocarditis, AF, antihypertensive drugs usage, antiarrhythmic drugs usage, and anticoagulation therapy (**Table 1**). However, statistically significant differences were observed for the following variables: age, CHF, diabetes mellitus (DM), pulmonary HTN, statin usage, use of antiplatelet agents, and use of DM drugs.

**Figure 1.**
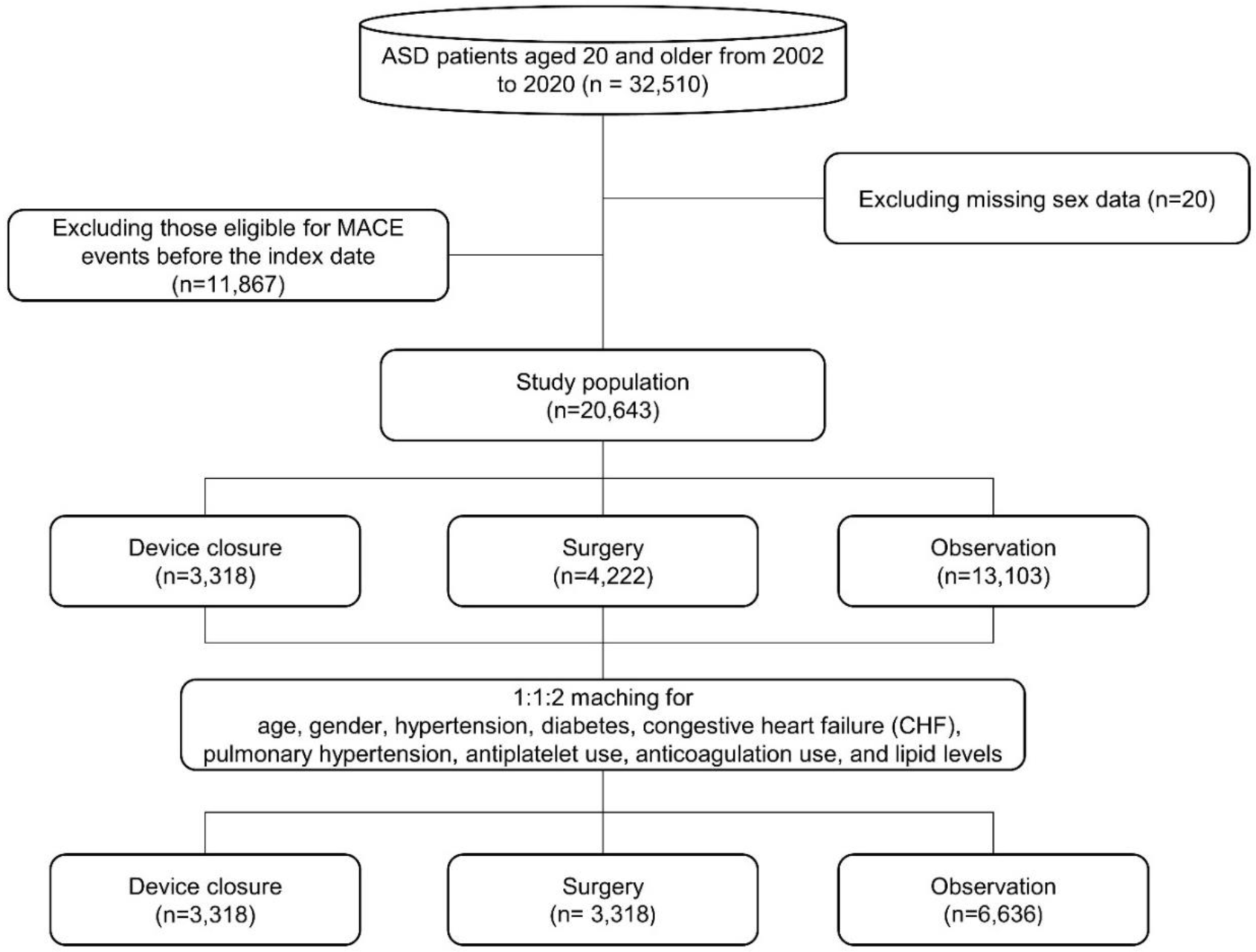
Study flow chart.

**Table 1.** Baseline characteristics.

|  | Before PSM |  |  |  |  | After PSM |  |  |  |  |
| --- | --- | --- | --- | --- | --- | --- | --- | --- | --- | --- |
|  | No | Surgery | Device | Total | p | No | Surgery | Device | Total | p |
|  | procedure |  | closure |  |  | procedure |  | closure |  |  |
|  | (N=13103) | (N=4222) | (N=3318) | (N=20643) |  | (N=6636) | (N=3318) | (N=3318) | (N=13272) |  |
| Male | 4630<br>(35.3%) | 1392 (33.0%) | 945<br>(28.5%) | 6967 (33.7%) | <0.001 | 1949<br>(29.4%) | 948 (28.6%) | 945 (28.5%) | 3842<br>(28.9%) | 0.561 |
| Age | 45.7 ± 16.7 | 44.3 ± 13.3 | 45.5 ± 13.1 | 45.4 ± 15.5 | <0.001 | 45.6 ± 13.1 | 44.3 ± 12.9 | 45.5 ± 13.1 | 45.3 ± 13.1 | <0.001 |
| CHF | 1400<br>(10.7%) | 1019 (24.1%) | 542<br>(16.3%) | 2961 (14.3%) | <0.001 | 997 (15.0%) | 590 (17.8%) | 542 (16.3%) | 2129<br>(16.0%) | 0.002 |
| DM | 2673<br>(20.4%) | 964 (22.8%) | 915<br>(27.6%) | 4552 (22.1%) | <0.001 | 1933<br>(29.1%) | 792 (23.9%) | 915 (27.6%) | 3640<br>(27.4%) | <0.001 |
| Hypertension | 3598<br>(27.5%) | 1672 (39.6%) | 1065<br>(32.1%) | 6335 (30.7%) | <0.001 | 2210<br>(33.3%) | 1064<br>(32.1%) | 1065<br>(32.1%) | 4339<br>(32.7%) | 0.325 |
| Pulmonary<br>hypertension | 202 (1.5%) | 362 (8.6%) | 189 (5.7%) | 753 (3.6%) | <0.001 | 192 (2.9%) | 215 (6.5%) | 189 (5.7%) | 596 (4.5%) | <0.001 |
| Endocarditis | 93 (0.7%) | 83 (2.0%) | 26 (0.8%) | 202 (1.0%) | <0.001 | 45 (0.7%) | 25 (0.8%) | 26 (0.8%) | 96 (0.7%) | 0.819 |
| Atrial<br>fibrillation | 765 (5.8%) | 547 (13.0%) | 244 (7.4%) | 1556 (7.5%) | <0.001 | 464 (7.0%) | 255 (7.7%) | 244 (7.4%) | 963 (7.3%) | 0.44 |
| Antihypertensive drugs | 4107<br>(31.3%) | 1910 (45.2%) | 1319<br>(39.8%) | 7336 (35.5%) | <0.001 | 2546<br>(38.4%) | 1352<br>(40.7%) | 1319<br>(39.8%) | 5217<br>(39.3%) | 0.06 |
| Statin | 1343<br>(10.2%) | 405 (9.6%) | 564<br>(17.0%) | 2312 (11.2%) | <0.001 | 1084<br>(16.3%) | 381 (11.5%) | 564 (17.0%) | 2029<br>(15.3%) | <0.001 |
| Antiplatelet | 1758<br>(13.4%) | 678 (16.1%) | 813<br>(24.5%) | 3249 (15.7%) | <0.001 | 1420<br>(21.4%) | 591 (17.8%) | 813 (24.5%) | 2824<br>(21.3%) | <0.001 |
| Antiarrhythmic drugs | 1569<br>(12.0%) | 725 (17.2%) | 472<br>(14.2%) | 2766 (13.4%) | <0.001 | 957 (14.4%) | 474 (14.3%) | 472 (14.2%) | 1903<br>(14.3%) | 0.961 |
| Anticoagulation | 354 (2.7%) | 200 (4.7%) | 111 (3.3%) | 665 (3.2%) | <0.001 | 209 (3.1%) | 112 (3.4%) | 111 (3.3%) | 432 (3.3%) | 0.789 |
| DM drugs | 639 (4.9%) | 166 (3.9%) | 176 (5.3%) | 981 (4.8%) | 0.011 | 445 (6.7%) | 142 (4.3%) | 176 (5.3%) | 763 (5.7%) | <0.001 |

Covariates for PSM: Age, sex, hypertension, diabetes, CHF, pulmonary hypertension, and medications (antiplatelet, anticoagulation, and lipid levels).
Abbreviations: PSM, propensity score matching; DM, diabetes mellitus; CHF, congestive heart failure

### Outcomes during long-term follow-up

The event-free survival and hazard ratios (HR) for MACEs during the 5-year follow-up period are shown in **Figure 2** and **Table 2**. We calculated crude HRs and further adjusted for covariates by including unadjusted variables in the PSM analysis. Regarding MACEs, compared with the observation group, the surgery group had the most favorable outcome, with an adjusted HR (aHR) of 0.72 (95% CI: 0.66–0.79). The device closure group demonstrated a better outcome than that of the observation group, albeit less favorable than that of the surgery group, with an aHR of 0.85 (95% CI: 0.78–0.92).

**Figure 2.**
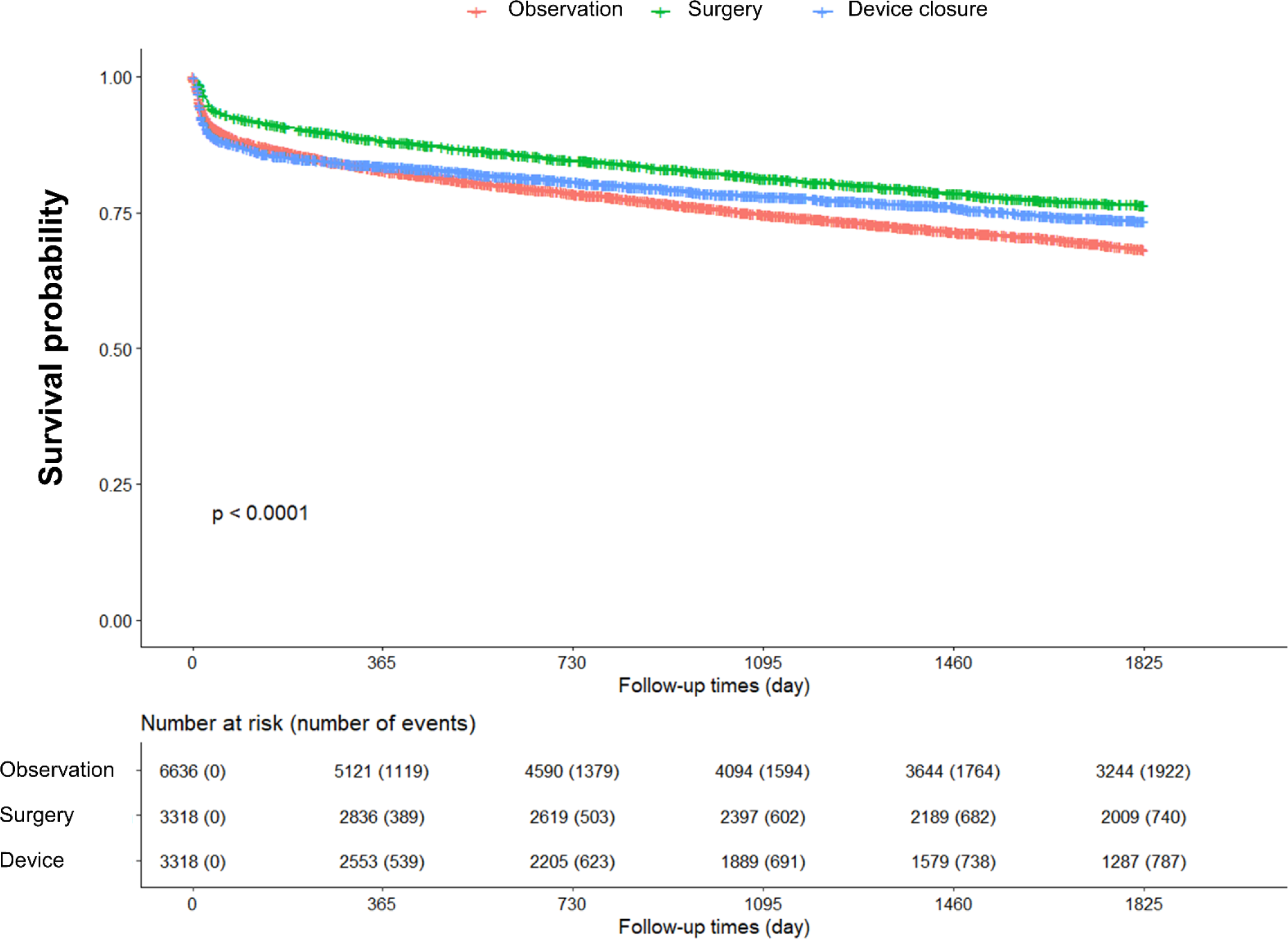
Five-year event-free survival curve for major adverse cardiovascular events.

**Table 2.**
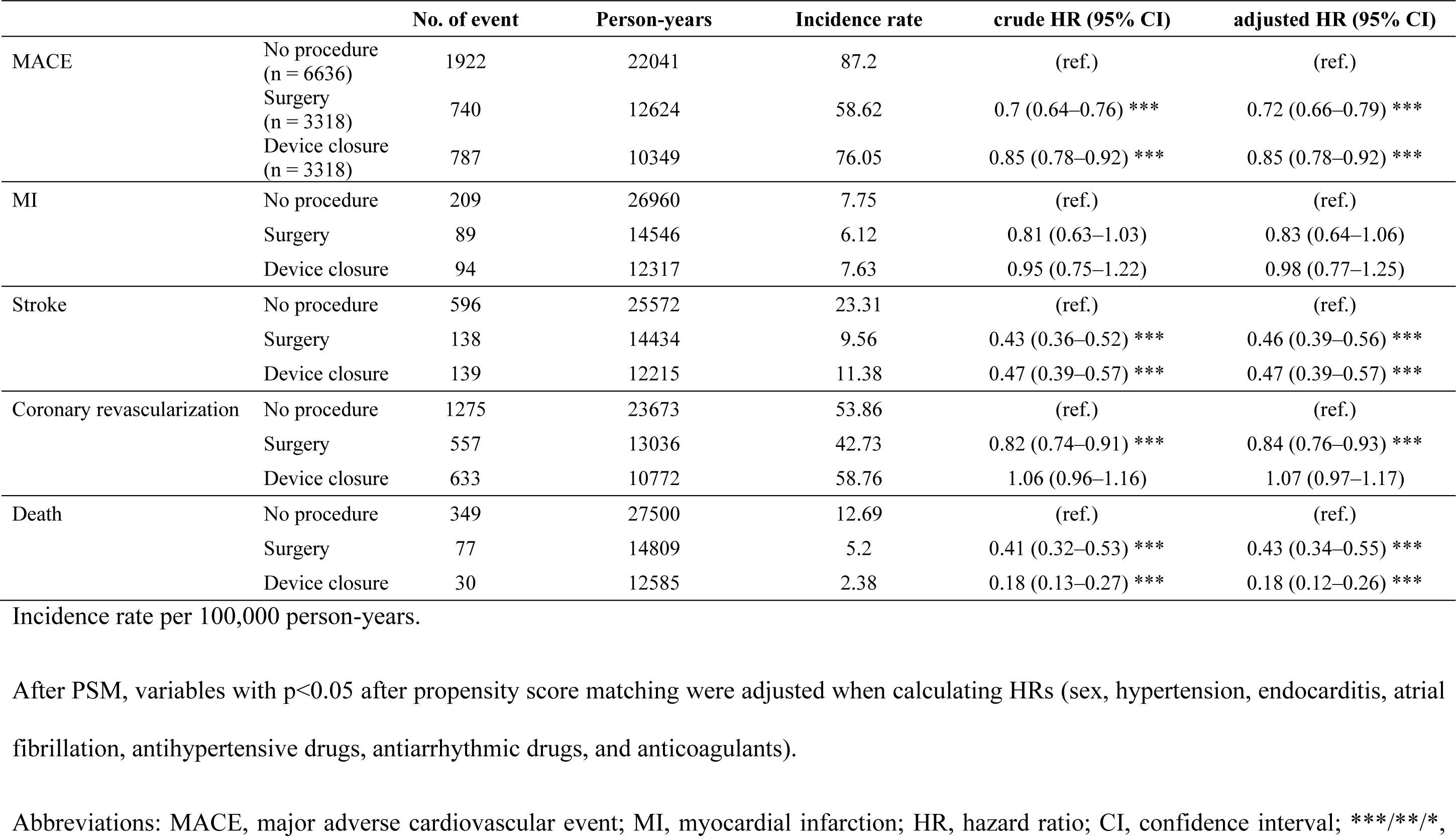
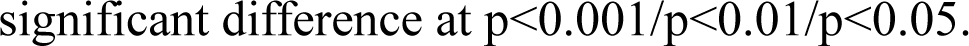
Hazard ratios for each cardiovascular event during the 5-year follow-up.

|  |  | No. of event | Person-years | Incidence rate | crude HR (95% CI) | adjusted HR (95% CI) |
| --- | --- | --- | --- | --- | --- | --- |
| MACE | No procedure<br>(n = 6636) | 1922 | 22041 | 87.2 | (ref.) | (ref.) |
|  | Surgery<br>(n = 3318) | 740 | 12624 | 58.62 | 0.7 (0.64–0.76) *** | 0.72 (0.66–0.79) *** |
|  | Device closure<br>(n = 3318) | 787 | 10349 | 76.05 | 0.85 (0.78–0.92) *** | 0.85 (0.78–0.92) *** |
| MI | No procedure | 209 | 26960 | 7.75 | (ref.) | (ref.) |
|  | Surgery | 89 | 14546 | 6.12 | 0.81 (0.63–1.03) | 0.83 (0.64–1.06) |
|  | Device closure | 94 | 12317 | 7.63 | 0.95 (0.75–1.22) | 0.98 (0.77–1.25) |
| Stroke | No procedure | 596 | 25572 | 23.31 | (ref.) | (ref.) |
|  | Surgery | 138 | 14434 | 9.56 | 0.43 (0.36–0.52) *** | 0.46 (0.39–0.56) *** |
|  | Device closure | 139 | 12215 | 11.38 | 0.47 (0.39–0.57) *** | 0.47 (0.39–0.57) *** |
| Coronary revascularization | No procedure | 1275 | 23673 | 53.86 | (ref.) | (ref.) |
|  | Surgery | 557 | 13036 | 42.73 | 0.82 (0.74–0.91) *** | 0.84 (0.76–0.93) *** |
|  | Device closure | 633 | 10772 | 58.76 | 1.06 (0.96–1.16) | 1.07 (0.97–1.17) |
| Death | No procedure | 349 | 27500 | 12.69 | (ref.) | (ref.) |
|  | Surgery | 77 | 14809 | 5.2 | 0.41 (0.32–0.53) *** | 0.43 (0.34–0.55) *** |
|  | Device closure | 30 | 12585 | 2.38 | 0.18 (0.13–0.27) *** | 0.18 (0.12–0.26) *** |

Incidence rate per 100,000 person-years.
After PSM, variables with $p < 0.05$ after propensity score matching were adjusted when calculating HRs (sex, hypertension, endocarditis, atrial fibrillation, antihypertensive drugs, antiarrhythmic drugs, and anticoagulants).
Abbreviations: MACE, major adverse cardiovascular event; MI, myocardial infarction; HR, hazard ratio; CI, confidence interval; \*\*\*/\*\*/\*,
significant difference at $p < 0.001$ / $p < 0.01$ / $p < 0.05$ .

**Figure 3** and **Table 2** show event-free survival and HRs for MI, coronary revascularization, stroke, and all-cause mortality. Surgery and device closure groups showed similar beneficial effects on stroke prevention. In contrast, for MI, there were no significant differences in the aHRs between the surgery/device closure and observation groups. For all-cause deaths, the device closure group had the lowest risk, followed by the surgery group and the observation group.

**Figure 3.**
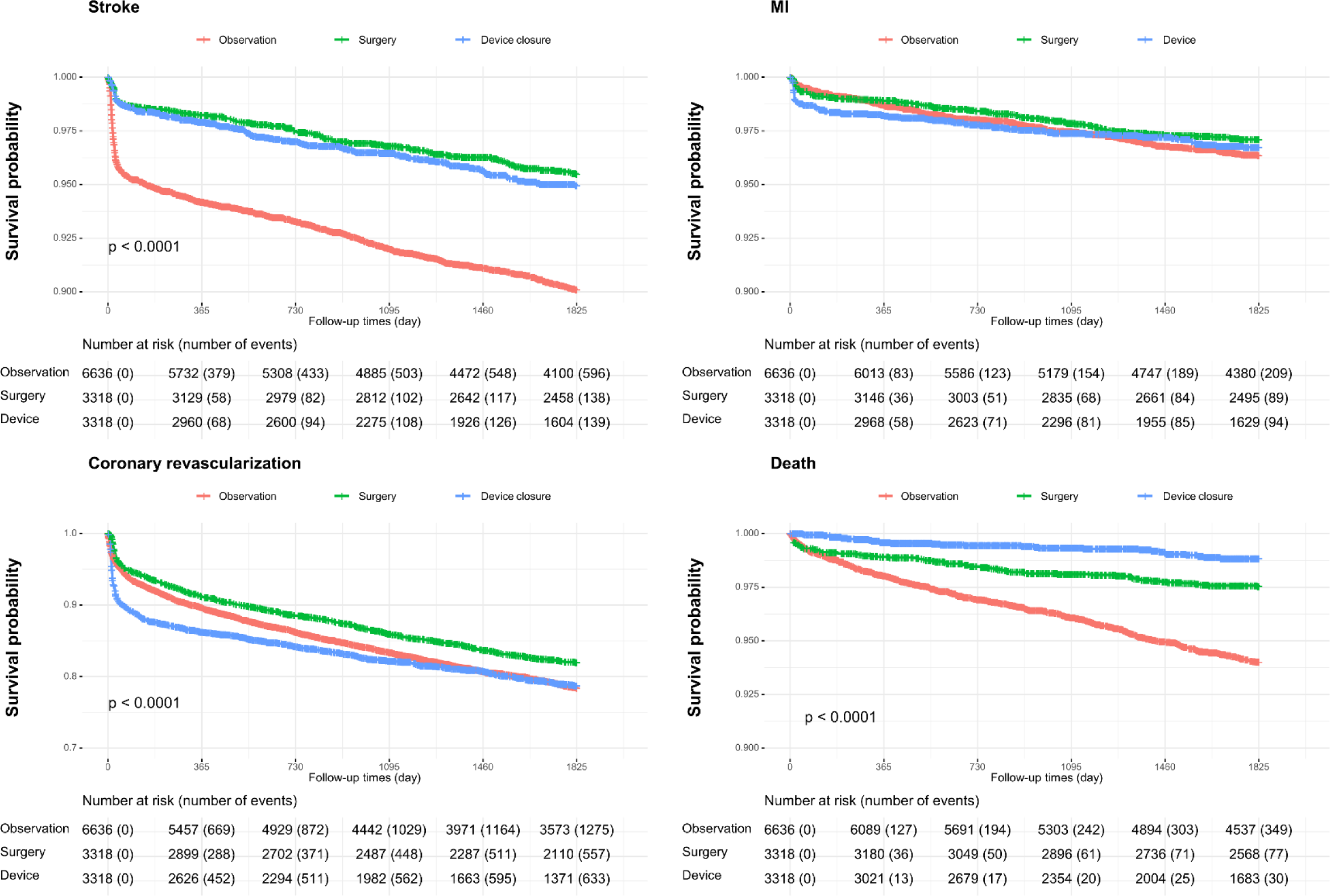
Event-free survival and hazard ratios for myocardial infarction, coronary revascularization, stroke, and all-cause mortality.

We also investigated the risk of stroke subtypes, including ischemic stroke and hemorrhagic stroke, as well as each component of hemorrhagic stroke, such as subarachnoid, intracerebral, and subdural hemorrhages, by comparing the surgery and device closure groups with the follow-up group (**Supplementary** Figure 1). Irrespective of the stroke subtype, the beneficial effect of both interventions was similarly sustained; however, this effect might be driven by a reduction in the occurrence of ischemic stroke. Among patients with hemorrhagic stroke, both the surgery and device closure groups had lower HRs for intracerebral hemorrhage than those in the observation group. However, for subarachnoid and subdural hemorrhages, no significant differences were observed between the groups.

### Post-procedural outcomes (surgery *vs*. device closure)

We investigated the post-procedural outcomes (until 6 months) after ASD closure in patients (**Table 3**). The occurrence of MACE was significantly higher in the device closure group than in the surgery group, with an aHR of 1.70 (95% CI: 1.45–2.00). MI (aHR: 1.68, 95% CI: 1.02–2.77) and coronary revascularization (adjusted HR 2.11, 95% CI 1.75-2.54) showed higher risks in the device closure, while mortality (aHR: 0.09, 95% CI: 0.02–0.36) was significantly lower in the device closure group than in the surgery group. No significant difference was observed between the two groups regarding stroke occurrence (aHR: 1.00, 95% CI: 0.65–1.53).

**Table 3.** Hazard ratios for major adverse cardiovascular events during the 6-month period following cardiac procedures.

|  |  | No. of event | Person-years | Incidence rate | crude HR (95% CI) | adjusted HR (95% CI) |
| --- | --- | --- | --- | --- | --- | --- |
| MACE | Surgery (n = 3318) | 244 | 632 | 386.08 | (ref.) | (ref.) |
|  | Device closure (n = 3318) | 407 | 600 | 678.33 | 1.75 (1.49–2.05) *** | 1.7 (1.45–2) *** |
| MI | Surgery | 25 | 656 | 38.11 | (ref.) | (ref.) |
|  | Device closure | 43 | 652 | 65.95 | 1.73 (1.05–2.83) * | 1.68 (1.02–2.77) * |
| Stroke | Surgery | 42 | 653 | 64.32 | (ref.) | (ref.) |
|  | Device closure | 44 | 652 | 67.48 | 1.05 (0.69–1.60) | 1 (0.65–1.53) |
| Coronary revascularization | Surgery | 166 | 639 | 259.78 | (ref.) | (ref.) |
|  | Device closure | 344 | 609 | 564.86 | 2.16 (1.8–2.6) *** | 2.11 (1.75–2.54) *** |
| Death | Surgery | 23 | 659 | 34.9 | (ref.) | (ref.) |
|  | Device closure | 2 | 658 | 3.04 | 0.09 (0.02–0.37) *** | 0.09 (0.02–0.36) *** |

Incidence rate per 100,000 person-years.
After PSM, variables with $p < 0.05$ after propensity score matching were adjusted when calculating HRs (sex, hypertension, endocarditis, atrial fibrillation, antihypertensive drugs, antiarrhythmic drugs, and anticoagulants).
Abbreviations: MACE, major adverse cardiovascular event; MI, myocardial infarction; HR, hazard ratio; CI, confidence interval; \*\*\*/\*\*/\*, significant difference at $p < 0.001/p < 0.01/p < 0.05$ .

### Subgroup analysis

A subgroup analysis of MACE in each group is shown in **Figure 4**. Overall, no significant differences were observed across each subgroup, and compared with device closure, surgery resulted in a lower HR for MACE.

**Figure 4.**
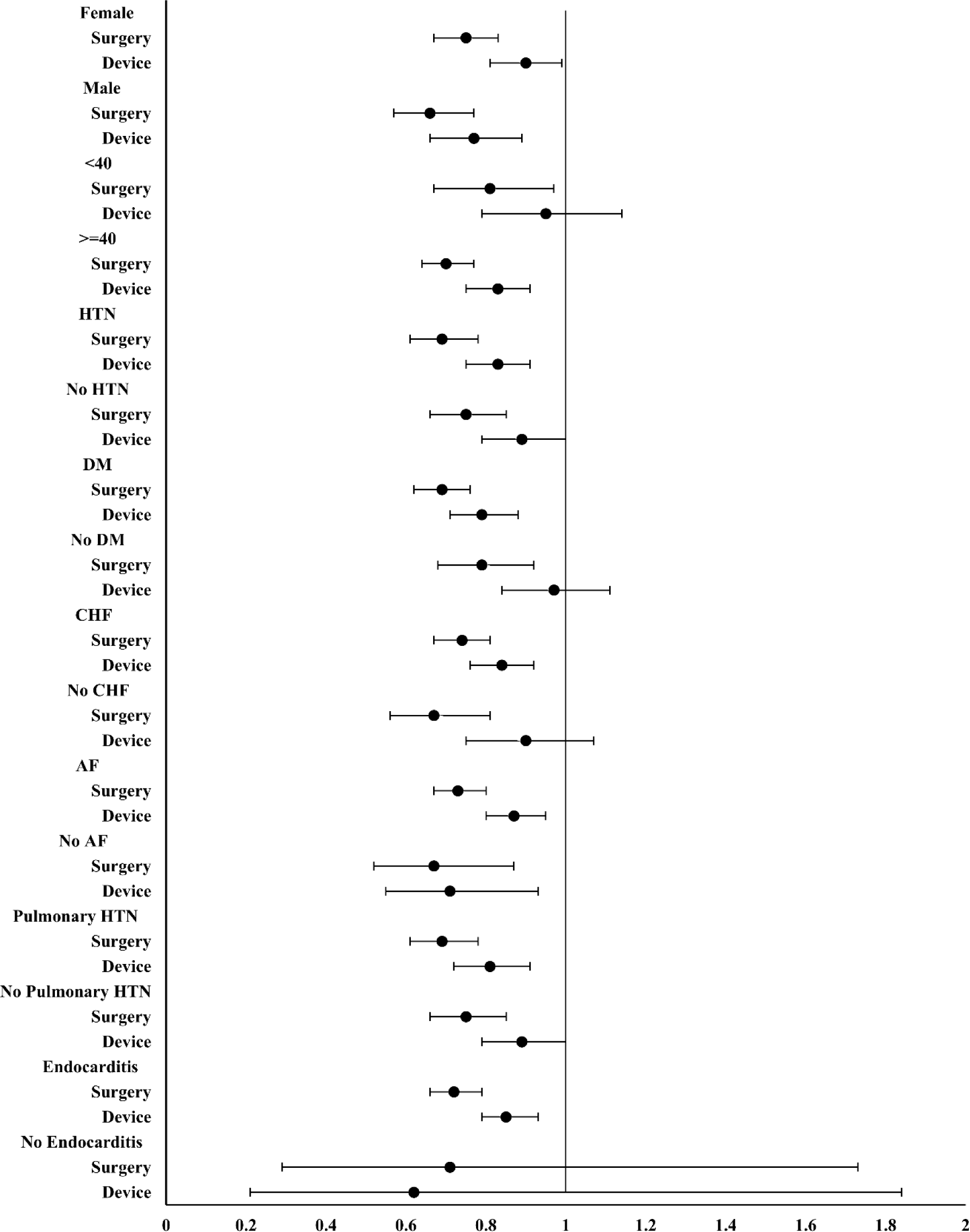
Subgroup analysis of hazard ratios for major adverse cardiovascular events.

## Discussion

We investigated the short- and long-term effects of different procedures on cardiovascular events in adults with ASD. Our findings revealed that, compared to no intervention, surgery exhibited a more beneficial effect in terms of MACE, followed by device closure. Moreover, the advantageous effect of surgery over device closure was maintained over the 6-month post-procedural period. However, the magnitude and direction of the beneficial effects vary according to specific vascular events. Specifically, device closure seemed more beneficial in terms of all-cause death, irrespective of the follow-up period, whereas surgical correction reduced the occurrence of coronary revascularization.

Individuals with ASD have a higher risk of cardiovascular complications when compared with the general population.^2,8,9^ Even after ASD closure, the prevalence of various cardiovascular complications was higher in individuals with ASD than in the general population.^10,13^ However, in cases where symptoms are present or there is progression in right ventricular remodeling or an increase in pulmonary arterial pressure, early ASD closure is recommended.^2,14^ Furthermore, in patients with ASD, undergoing ASD closure has been associated with a lower incidence of AF, stroke, and mortality compared with cases where closure was not performed.^13^ Some studies have reported no difference in cardiovascular outcomes among patients with ASD irrespective of intervention.^8,9,15^ However, other studies have reported superior outcomes in the intervention group compared with the surgical group in terms of complications including stroke and mortality.^11,13,16,17^

Cardiac disease, with heart failure as the most prevalent, accounts for nearly half of the causes of mortality in patients who do not undergo ASD closure.^9^ However, studies investigating the risk of various stroke subtypes, ischemic heart disease, and other cardiovascular outcomes in adult patients with ASD are lacking.

In this study, we classified adult patients with ASD into different treatment groups, compared cardiovascular outcomes over a 5-year period, and assessed post-procedural outcomes within 6 months in those who underwent ASD closure.

The overall outcomes, including MACE, were more favorable in ASD closure groups than in the observation group. When each outcome was examined individually, the prognosis was better in the ASD closure groups than in the observation group. The most direct cause for this may be the prevention of paradoxical embolism due to ASD closure.^1,6,18^ Additionally, ASD closure may lead to the recovery of cardiac volume overload and tissue remodeling caused by ASD, delay the progression to heart failure, and reduce the possibility of cardiovascular events.^1,5,18^ Furthermore, this study found that the beneficial effect of ASD closure, irrespective of the intervention type, is more prominent in the cerebrovascular bed than in the cardiovascular bed after adjusting for several relevant factors including HTN, antithrombotic medication usage, and statin usage. While it is difficult to determine the reason for the effectiveness of ASD closure in preventing cerebrovascular diseases over other cardiovascular conditions, one hypothesis is that paradoxical thromboembolism may have a greater impact on cerebral vessels than on coronary arteries. Additionally, although the mechanism by which ASD closure reduces the risk of cerebral hemorrhage is not entirely clear, chronic changes in the cerebrovascular bed due to ASD possibly increase vulnerability to cerebral hemorrhage, and these changes may be reversed following ASD closure. Future studies with more rigorous design are required to determine the mechanisms underlying this phenomenon.

Furthermore, within the ASD closure group, the surgery group yielded superior results when compared to the device closure group. However, caution should be exercised when conclusively asserting that surgery yields better outcomes than those of device closure. Examining the event numbers for each outcome, it is evident that in both the short and long-term period, coronary revascularization contributed to more than half of the overall MACE. Therefore, it can be postulated that the impact of coronary revascularization plays a pivotal role in the overall outcomes of MACE. Providing a rational explanation for the increased frequency of coronary revascularization in patients who underwent device closure compared with those who underwent surgical closure is also challenging. However, it can be hypothesized that the thrombotic risk associated with the device itself may be higher than that associated with surgery. Additionally, the increased frequency of coronary angiography performed during device closure procedures could be a contributing factor. Each outcome demonstrated differences in prognosis based on treatment approach, underscoring the need to carefully consider these factors.

### Limitations

This study has a few limitations. First, there is an inevitable possibility of inaccuracies or errors in the data provided because this study was based on data from the NHIS. Second, although we tried to reduce the imbalance between the groups through PSM, we could not consider other important factors, such as the type, size, or shunt degree of the ASD, detailed information on the catheter or surgical closure (device type or surgical technique), and lifestyle habits relevant to vascular risk factors, such as alcohol consumption, smoking, and physical activity. Third, national claims data may have a margin of error of approximately 2 weeks due to discrepancies between the actual procedure date and the billing date. To reduce this time gap, we examined events over a 6-month period when assessing short-term outcomes.

### Conclusions

ASD closure, whether by surgery or using a device, is associated with a decreased incidence of cardiovascular outcomes in adults. Moreover, the benefits on cardiovascular outcomes vary with the type of closure method, underscoring the need for a tailored approach to manage ASD in adults.

## Data Availability

The data presented in this study are not publicly available, but can be made available upon reasonable request to the corresponding author and with permission from the Korea National Health Insurance Service.

## Acknowledgements

None

